# GMRC: Gene Median Ratio Clustering Algorithm to Rank Clusters and Identify Driver Genes in Intrahepatic Cholangiocarcinoma

**DOI:** 10.1101/2022.11.16.22282432

**Authors:** Nandita Puri, Ridhi Arora, Vishesh Kumar Tanwar, Sajal K. Das

**Author notes:** [equal contribution]. {, }.

## Abstract

Intrahepatic Cholangiocarcinoma (iCCA) is an asymptomatic malignancy of the bile ducts in the liver. The research is predicated on the assumption that a disease is a cluster of genes interacting in a regulatory network and a few driver genes regulating the network. Keeping this in mind, the paper proposes a Gene Median Ratio Clustering (GMRC) algorithm to independently rank the clusters and identify driver genes by hybridizing *gene-median ratio* (GMR) and *gene cluster network* (GCN). GMR then employs the Poisson distribution to translate right-skewed data into the Normal distribution and the Anscombe transformation to eliminate data noise and determine the median difference in gene concentration between distinct disease stages. Additionally, hierarchical clustering is separately applied to raw data for gene clustering based on the RNA sequence count. In the process, GMRC is evaluated over the GSE32225 dataset of 155 patients to extract 12 clusters network for proliferation patients.

## 1 Introduction

Intrahepatic Cholangiocarcinoma (iCCA) is one of the epithelial malignancies that fall within the metastatic group of heterogeneous malignant biliary neoplasms. The worrisome mortality of these tumors, which accounts for around 10% of all cancer-related fatalities globally each year, results from their stealthy appearance, high aggressiveness, and resistance to treatment (Buettner et al. 2017). Pathologists detect iCCA incidentally in 0.5–7.4% of autopsies and 1–3% of cholecystectomy samples. Current non-invasive methods for diagnosing iCCA are inadequate, and histological confirmation is essential. High heterogeneity of iCCA at the genomic, epigenetic, and molecular levels significantly reduces the efficacy of present therapies.

The research is motivated by the need for a clear distinction between key and driver genes. Key genes are those with the most significant *log2fold* change and are the primary nodes of a protein-to-protein network, while driver genes are those that accelerate or inhibit the disease-causing cluster. The genes communicate with one another to raise the transcription rate, forming a disease gene regulatory system. In a gene cluster network (GCN), driver genes either promote or inhibit the whole disease regulatory system. Researchers have discovered numerous potential biomarkers; however, only a few have been discovered and classified to identify driver genes without using protein-to-protein interactions. Researchers have identified key genes using the *log2fold* algorithm with tight cutoffs. Not all these genes are potential biomarkers, but they can be mediating genes, meaning they are not oncogenes but communicate with driver genes or immune-related genes to promote other activities, such as cell proliferation or metabolism. Fang *et al*. (Fang et al. 2021) modeled the data into normal distribution and conducted variance transformation to account for differential sequencing depth as a technical error. Tian *et al*. (Tian et al. 2019) deployed the hierarchical clustering algorithm to cluster genes and utilized phenotype traits such as age, gender, and height to rank clusters and identify driver genes. However, existing methodologies hardens the task of identifying driver genes or disease gene cluster network as phenotype traits may not be available in every clinical scenario.

Contrasting existing schemes, our proposed methodology, namely, Gene Median Ratio Clustering (GMRC), feeds the data in two independent processes: i) GMR a difference in the median ratio of a gene in distinct disease stages to identify overexpressed genes and shortlists genes with more than 100% median difference and ii) GCN - incorporation of the hierarchical clustering to cluster genes based on RNA-seq count. Upon receiving the desired findings (genes/biomarkers), GMRC ranks the clusters based on the number of common genes in GMR and GCN. Our core contributions are:

1. The proposed methodology GMRC independently employs the GMR and GCN such that the output of one algorithm does not confluence the other.
2. GMRC can rank and identify driver genes without any phenotypic trait or external protein-to-protein network. GMR deploys median instead of mean, immunizing the median ratio against high variations.

## 2 Proposed Scheme

This section discusses the data acquisition process and the details of our proposed scheme, GMRC.

### 2.1 Data Acquisition

We leveraged the GSE32225 (NCBI 2017) data collected from the NCBI website containing iCCA information of 155 patients. Out of 155, 6 patients belong to the *Normal* control group (no cancer), 92 and 57 are at the *Inflammation* and *Proliferation* stages in their bile duct tubes. There are around 24526 unique genes, and their RNA sequence counts in various disease stages of different patients have been reported.

### 2.2 Gene Median Ratio (GMR) Algorithm

The function of an organ or tissue influences the activation of genes for specific protein synthesis. A gene is overexpressed if there is a statistically significant difference or change between diseased and healthy individuals in the read counts or expression levels of RNA sequencing. The GMR algorithm aims to establish overexpressed oncogenes in proliferation disease-stage patients against the count of inflammation genes. Fig. 1 presents the schematic representation of our proposed methodology, and the details are given below:

**Figure 1:**
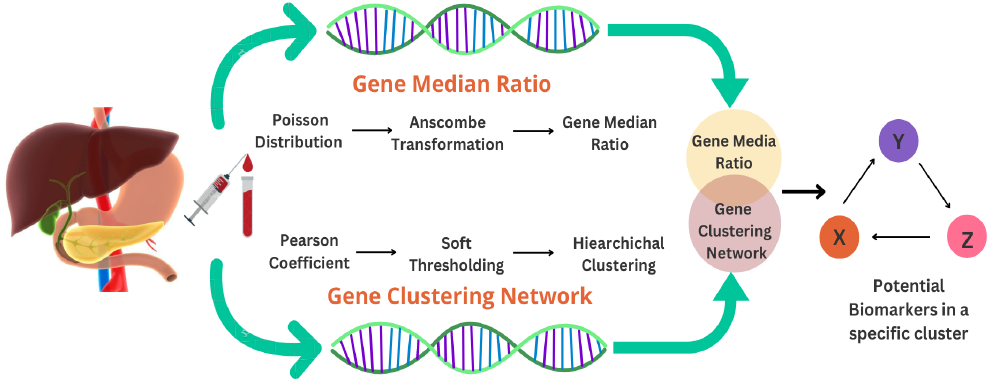
The schematic representation of our proposed methodology to rank clusters and identify driver genes.

1. **Outlier Detection** Outlier is a significant phase in RNA sequence data modeling because when a patient’s RNA sequence is extracted, the patient may have additional phenotypic traits,, *e*.*g*., drinking and smoking, that may increase the gene expression compared to the patient with no drinking/smoking history. We also deploy outlier detection to remove the variance from the data, such that mean and standard deviation are correlated. Addressing this concern, our proposed methodology utilizes principal component analysis (PCA) to extract data clusters and remove patients with excessive RNA-seq counts. For instance, the outliers, namely CCNY051, CCNY027, & CCNY012, in GSE32225 dataset are shown in Fig. 2.
2. **Poisson Distribution** Upon data modeling, it is identified that the data is not normally distributed and is highly right-skewed, as shown in Fig. 3 (left-side). For discrete data, sparse data makes it hard to forecast gene correlations or the chance of overexpressing a gene at a specific disease stage, inflammation, or proliferation. It is important to note that the Poisson distribution can only be deployed with correlated mean and standard deviation. Thus, we transformed each RNA count say *x*, as shown in Fig.3 (right-side) using the Poisson distribution, 𝒫 𝒟, with hyper-parameter *λ* = 10, as defined in Eq. **??**:

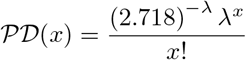
3. **Sequencing Depth: Anscombe Transformation** Biological replicates have comparable sequence counts and have the same transcriptional profile in a specific disease stage, either inflammation or proliferation. However, in most cases, the sequencing depth within a group will be similar and diverse due to technical errors and the nonavailability of phonetic metadata, which behaves as noise and significantly impact the biomarker extraction. Therefore, we employed the Anscombes’ transformation to remove such data noise from RNA sequence count, say *x*,

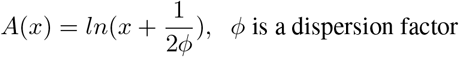
4. **Ratio Computation** The algorithm then determines median gene differences between disease phases, for instance, Gene A’s median in inflammation and proliferation. The median difference gives a proportion of gene overexpressed during proliferation, as shown in Fig. 4, where the vertical axis shows the gene levels.

**Figure 2:**
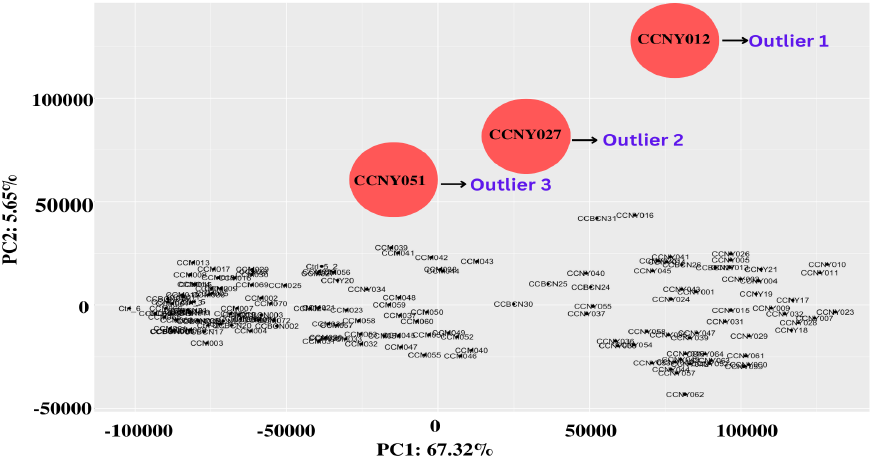
Detecting outlier patients in raw data using PCA.

**Figure 3:**
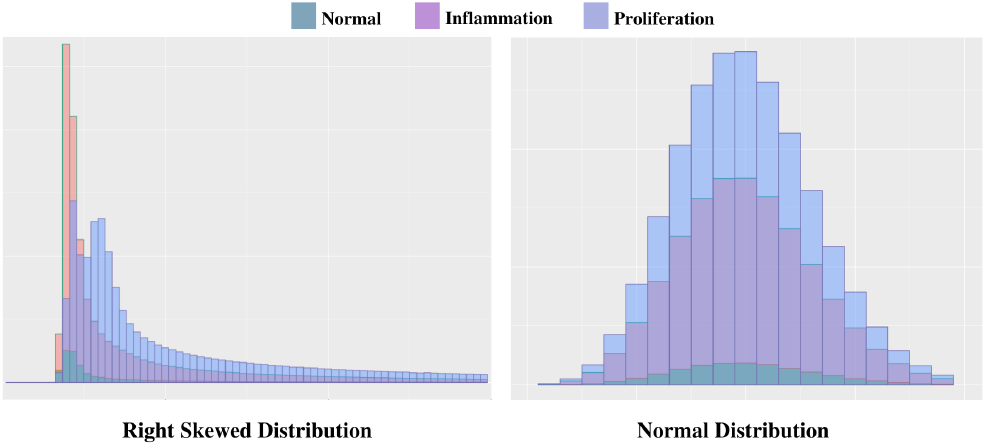
Normalized data distribution (right) with mean=10 of the right-skewed (left) raw data using the Poisson distribution.

**Figure 4:**
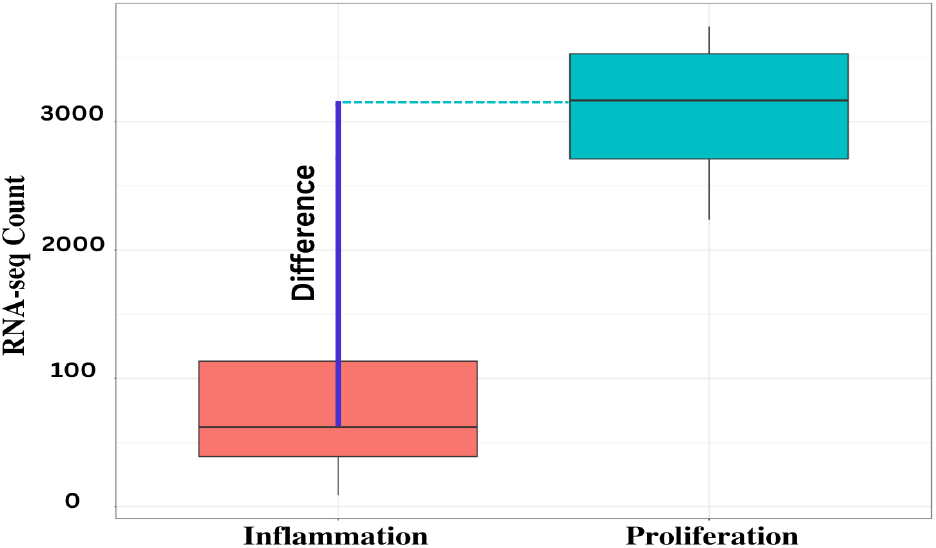
Median ratio to identify overexpressed genes in proliferation against inflammation.

### 2.3 Gene Clustering Network (GCN)

The interactions between genes in a network can lead to disease-causing protein production. As an example of a fundamental GCN, as depicted in Fig. 5. Gene X may generate a protein that activates Gene Y, which in turn produces a protein that activates Gene Z. Thus, a gene regulatory network is developed as a collection of molecular regulators that interact with other genes, proteins, or miRNA, forming a chain in the cell to regulate the gene expression levels of miRNA and proteins, controlling cell’s functioning. The detailed step-by-step working of GCN is summarised below.

**Figure 5:**
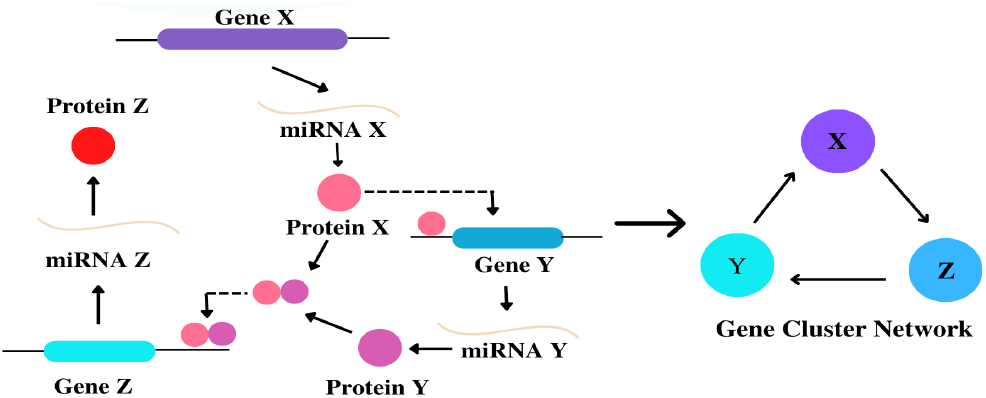
Gene cluster network depicting genes interaction, proteins formation to regulate the disease network.

1. **Pearson coefficient:** It is a method for calculating correlation coefficients that evaluate the strength of the association between two continuous/discrete variables. To quantify the association, a linear relationship is established between miRNA Gene X and Gene Y expression levels in their respective cell groups. If the data points are closer to the trend line, it infers Genes X and Y have a strong relationship. Therefore, if all data points lie on the trend line, the correlation equals 1, defined below.

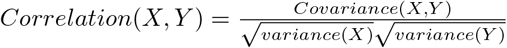
2. **Soft Thresholding:** After establishing pair-wise correlations between the genes, an adjacency matrix is generated. Soft thresholding (Zhang and Horvath 2005) emphasizes substantial connection (higher correlation coefficients) and suppresses low correlation (noise) by increasing the correlation’s strength. The RNA sequence is raised to a power that yields a scale-free network with power-law-distributed node connections. Most nodes have few connections, while a few hubs have many. This technique is performed on each power term until the adjacency matrix-derived graph network has a scale-free topology as depicted in Fig. 6, where the correlation values are raised to a power to obtain a scale-free topology network with soft thresholding (with a threshold value 6).
3. **Dissimilarity Measurement:** Distance between genes in the weighted adjacency matrix minus one is used to measure the similarity between the genes. To assess the topological overlap of Gene 1 and Gene 3 (ref. Fig. 6) with a similarity matrix of a threshold value, the rowand column-wise correlation are aggregated followed by dividing by the minimum of row/column-wise aggregation, and added one. Further, subtract one from the resulting value to obtain dissimilarity. High correlation means a higher topological overlap and lesser dissimilarity.
4. **Linkage Hierarchical Clustering:** Upon successfully computing the correlation and dissimilarity measure, we deploy hierarchical clustering to generate groups of similar genes having similar characteristics. Each data point is assigned to a separate cluster, compute the distance between each cluster, join the two most similar clusters, and disjoin the highly dissimilar gene. GMRC employs mean linkage as the distance metric, defined as the distance between each cluster’s point and other clusters.

**Figure 6:**
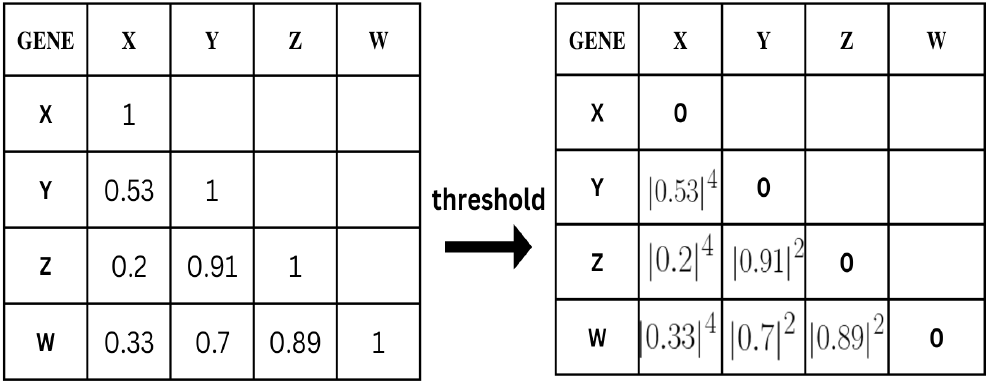
Soft thresholding to obtain a scale-free topological network.

### 2.4 Ranking Clusters and Identify Driver Genes

In the last phase of the GMRC algorithm, we rank the clusters and identify the driver genes in the clusters. The cluster ranking is as follows: the cluster with the highest number of identical genes in the GMR up-regulated gene series ranks the highest. The identical genes in both algorithms are the driver genes communicating with each other, and the remaining genes in the cluster are the mediating genes.

## 3 Results and Experimental Analysis

### 3.1 Gene Cluster Network Cluster Extraction

We only performed our experimental analysis for patients at the proliferation stage and observed that the data was partitioned into two distinct groups. We utilized hierarchical clustering and PCA, which concluded that there is a transitional time between inflammation and proliferation when patients migrate from the inflammation to the proliferation stage. Using the hierarchical clustering algorithm, 12 gene cluster networks are obtained, as tabulated in Table 1. Following the execution of the clustering method, two dendrograms comprising separate clusters are generated. Figs. 7(a) and (b) depict the unmerged and merged clusters, respectively. It can be observed that the turquoise cluster is the dominant cluster (by size), followed by the blue cluster.

**Table 1:** Extracted clusters using GMRC.

| Module Color | Genes | Module Color | Genes | Module Color | Genes |
| --- | --- | --- | --- | --- | --- |
| Black | 142 | Blue | 2952 | Brown | 2250 |
| Green | 178 | Green-Yellow | 26 | Grey | 16588 |
| Magenta | 67 | Pink | 91 | Purple | 27 |
| Red | 153 | Turquoise | 5064 | Yellow | 480 |

**Figure 7:**
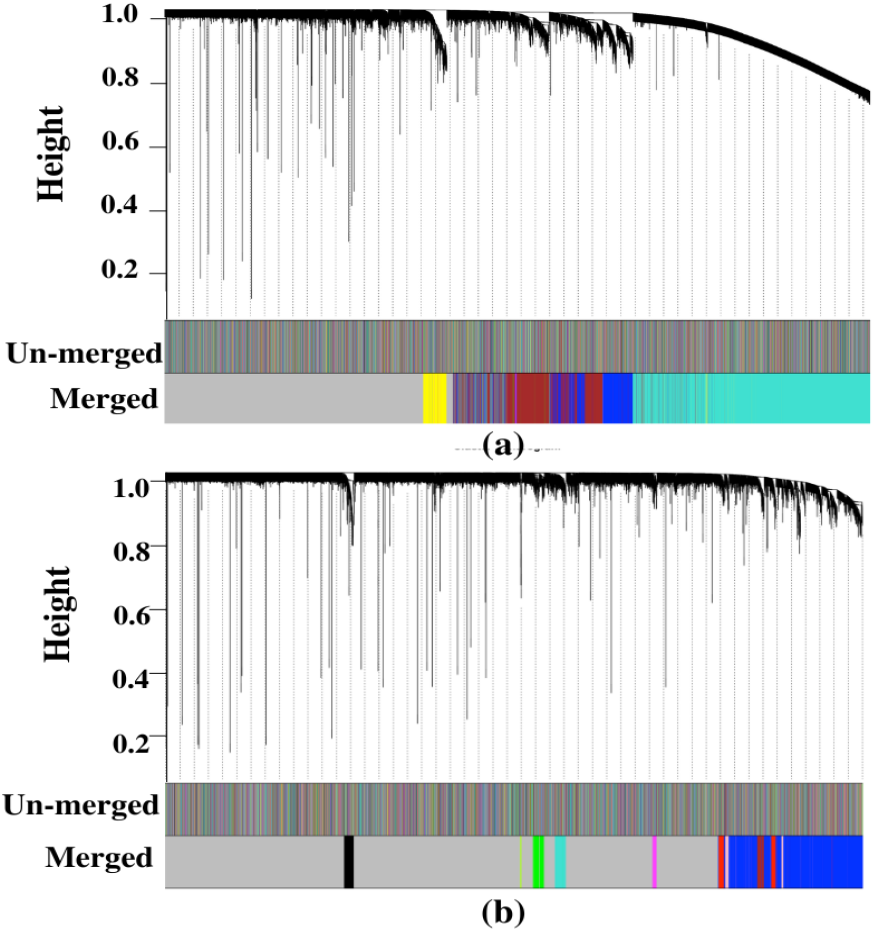
Hierarchical Clustering for Proliferation disease stage

### 3.2 Key Driver Genes

After obtaining the clusters in Section 3.1, GMRC ranks the clusters in the probability of being a disease-causing cluster. Thus, the brown cluster (refer Fig. 7 (a)) topped the rank with 2250 genes having 30 common genes with GMR. From 30, 22 genes are identified as driver genes using prior literature, as depicted in Table 2. These 22 genes are potential biomarkers in other carcinomas also. TUBA1B gene is overexpressed by 132% in the proliferation disease patients and is a potential biomarker in Wills Tumor (Xu et al. 2020), also known as nephroblastoma in pediatric patients. While being a potential biomarker in children’s kidney cancer, it is a potential biomarker in Hepatocellular Carcinoma, the same as CISD1 (Lu et al. 2022). CISD1 is overexpressed by 115% in our dataset in proliferation and is known to regulate ferroptosis negatively. Apart from this, SSU72 gene (Kim et al. 2022), which is overexpressed by 125%, is a potential biomarker in liver inflammation, leading to proliferation.

**Table 2:** Extracted Genes from the brown cluster (refer Fig. 7(a)) using GMRC algorithm.

| Biomarker | Gene Name | Biomarker | Gene Name |
| --- | --- | --- | --- |
| Hepatocellular | TUBA1B, SSU72, CISD1 | Ovarian | CCNY, ZFP36L1 |
| Breast | SEPTIN2, PAPOLA, BIRC2, NUSAP1, ATP6V1G1, PTGES3 | Colorectal Carcinoma | GID8, RAB7 |
| Gastric | RHOA, PCBP2, STK3 | Prostrate | TMEM256 |
| Gliomas | MTPN | Tumor Progression | CCZ1B, ITGB1 |
| Regulates Cancer Metasis | CD44 | Endometrial Carcinoma | YTHDC1 |

Contrasting, the researchers also observed a few genes which are potential biomarkers in other cancer types. For instance, SEPTIN2 (He et al. 2019) is a tumor belonging to the SEPT family of genes that increase the cellular process of hepatoma, glioblastoma, and mesangial carcinoma cells and are overexpressed by 131% in breast and biliary tract cancer. PAPOLA (Komini et al. 2021) is observed to be associated with proliferation, which is an ideal candidate target from prior research as it is overexpressed by 119% in triple-negative breast tumors. Likewise, BIRC2 (Samanta et al. 2020), NUSAP1 (De Luca, Romano, and Bucci 2021), ATP6V1G1 (Qiu et al. 2021), PTGES3 (Adekeye et al. 2022), that are overexpressed by 118%, 111%, 117%, and 118% respectively, are potential biomarkers in breast cancer, have also been overexpressed in iCCA proliferation, giving the researchers an opportunity for drug re-purposing. CCNY (Liu et al. 2016), and ZFP36L1 (Suk et al. 2018) are potential biomarkers in Ovarian cancer and are also overexpressed by 127% and 123%, respectively. Other potential biomarkers are RHOA (Nam, Kim, and Lee 2019), PCBP2 (Chen et al. 2018), and STK3 (Chen et al. 2021), which are found to be overexpressed by 112%, 120%, and 117%, respectively, and are found in Gastric carcinoma. It (Harmouch 2022) is caused by a germ called Helicobacter pylori which further causes a mutation in these genes. The last potential biomarkers obtained in our experiments in Colorectal Cancer genes are GID8 (Lu et al. 2017), RAB7 (Hu et al. 2021), which are highly overexpressed in iCCA proliferation by 110% and 112%, respectively.

## 4 Conclusion

This paper proposes a neoteric and independent process, GMRC, that integrates GMR and GCN, devoid of any clinical phenotypic feature. GMRC resists the varying results from technical mistakes or other diseases and does not need external data to rank clusters and identify oncogenes. Compared to existing methods that need an external clinical phenotype and PPI network to generate oncogenes, the suggested procedure produced 73% pure cancer-driving genes. The extracted oncogenes are TUBAB1B, SSU72, CISD1, SEPTIN2, PAPOLA, BIRC2, NUSAP1, ATP6V1G1, PGTES3, RHOA, PCBP2, STK3, CCNY, ZFP36L1, GID8, RAB7, CCZ1B, ITGB1, MTPN, CD44, YTHDC1, TMEM256. In addition, we can effectively extract a more significant number of possible oncogenes than conventional methods. These genes are potential indicators for other carcinomas, enabling more investigation into drug repurposing for rare diseases.

## Supporting information

Gene Distribution in various disease stages

## Data Availability

All data produced are available online at https://www.ncbi.nlm.nih.gov/geo/query/acc.cgi?acc=GSE32225

