## Supplementary material for "GMRC: Gene Median Ratio Clustering Algorithm to Rank Clusters and Identify Driver Genes in Intrahepatic Cholangiocarcinoma": Gene Distribution in various disease stages

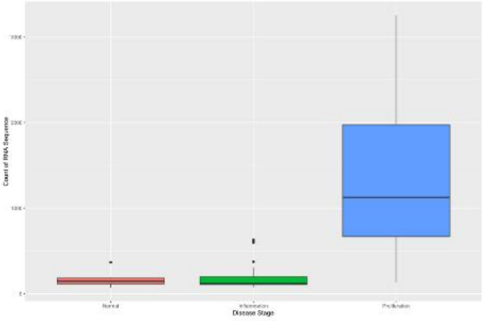


ATP6V1G1

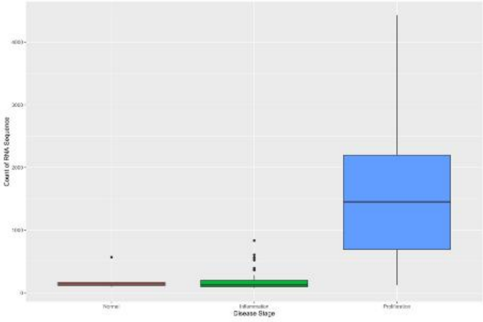


BIRC2

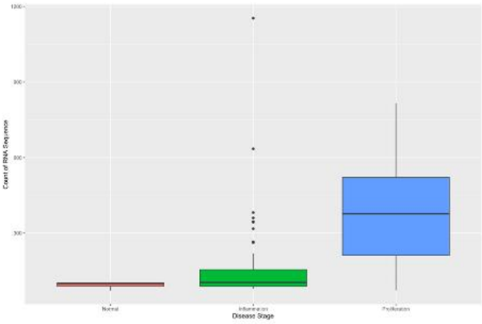


CCNY

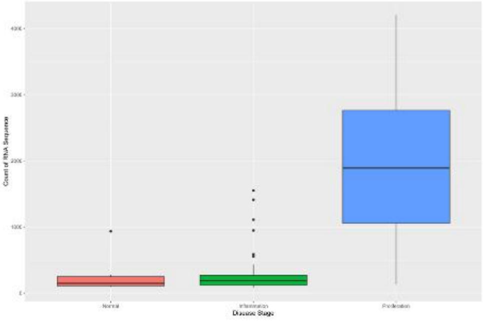


CCZ1B

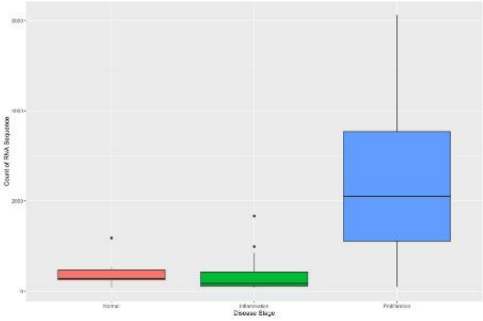


CD44

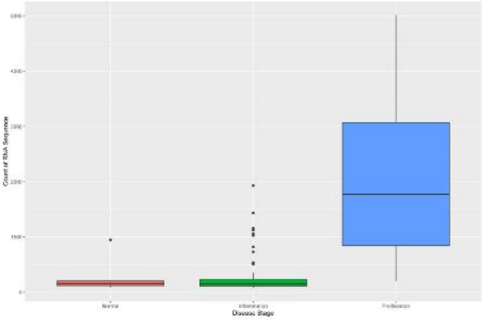


CISD1

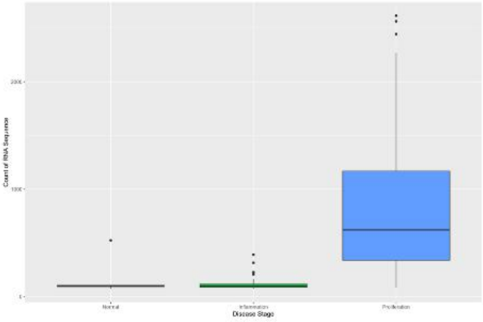


COL6A3

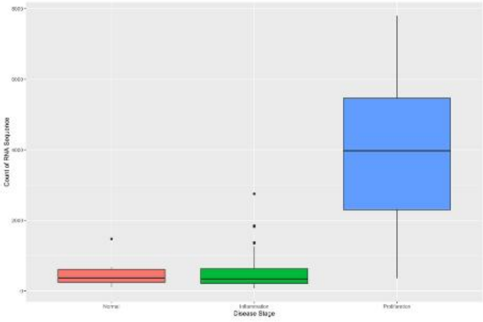


GID8

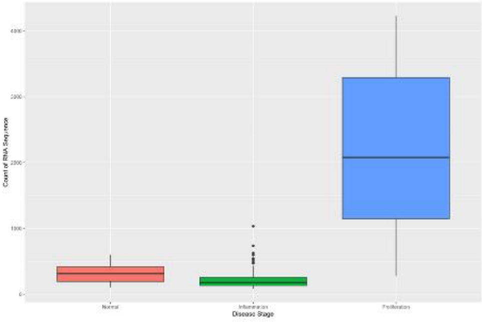


ITGB1

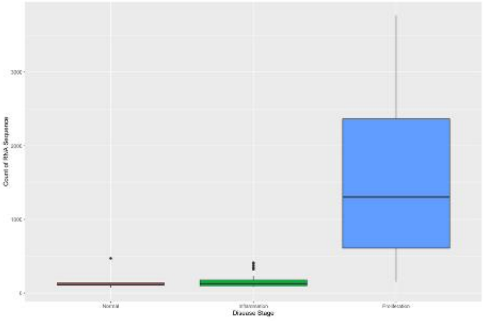


MTPN

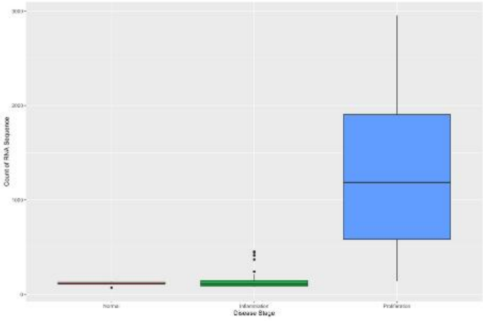


NARS1

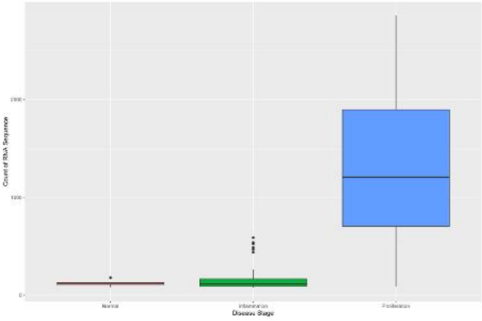


NDEL1

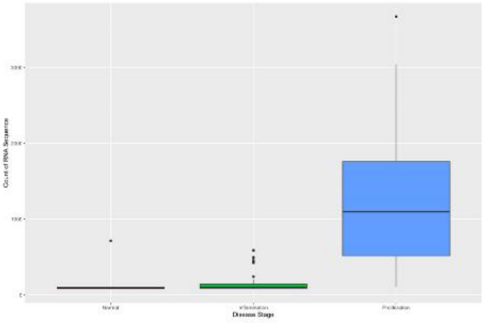


NUSAP

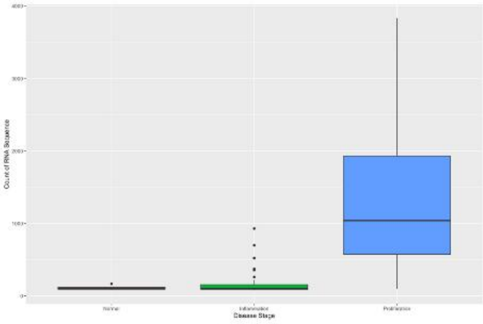


OXR1

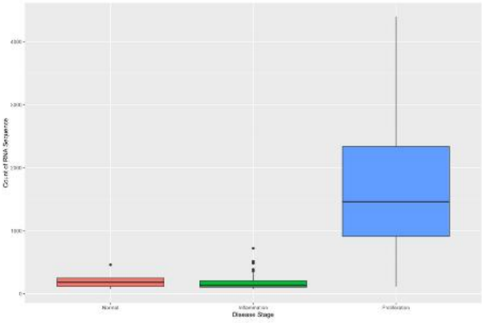


PAPOLA

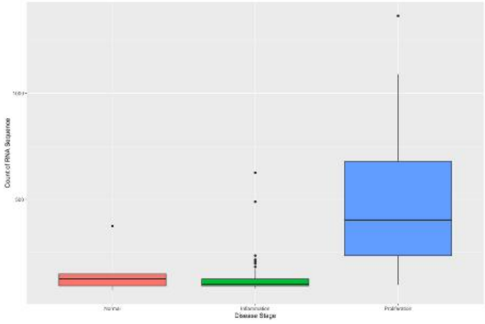

PCBP2

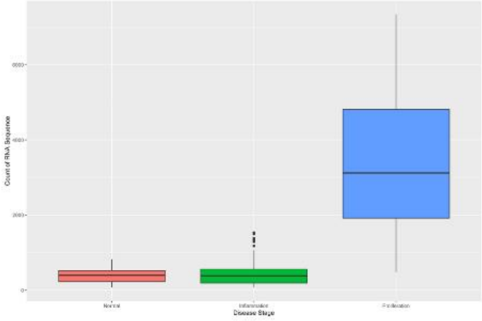

PSMA6

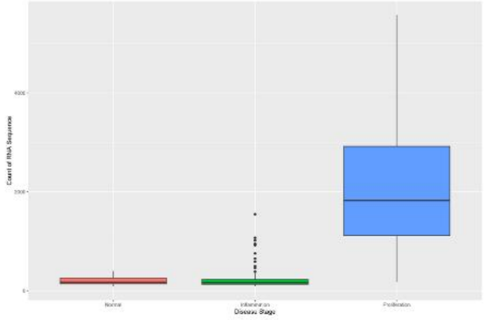

PTGES3

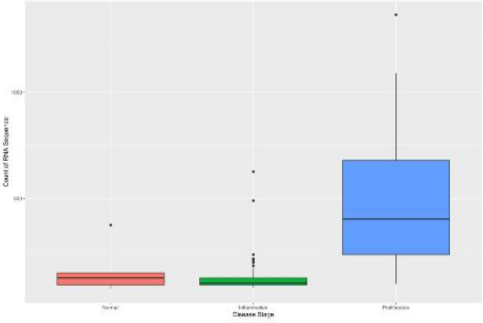

PTGES3

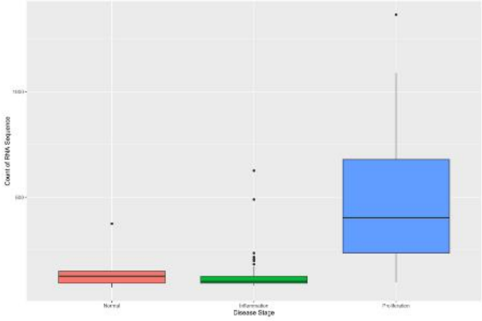

RAB7A

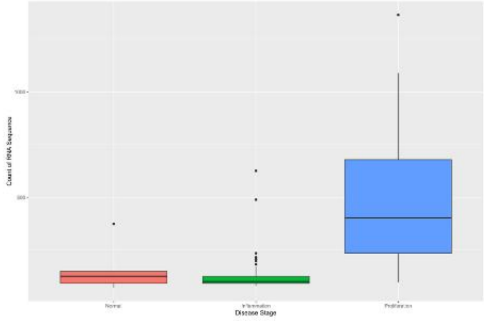

RHOA

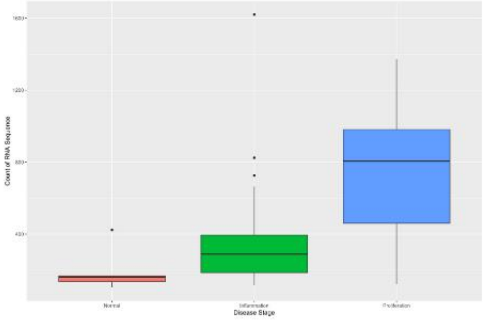

SEPTIN2

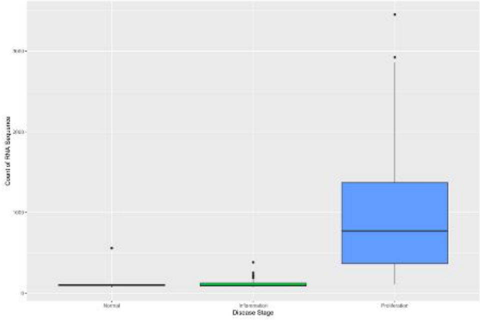

SLC7A1

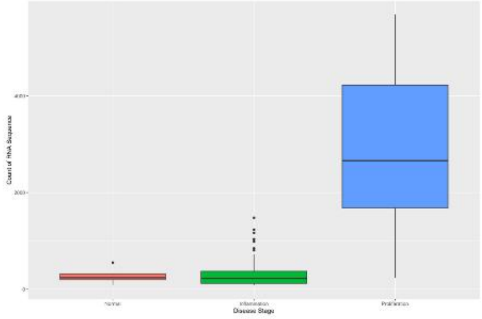

SSU72

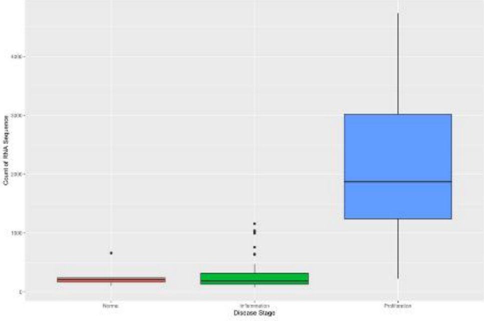

STK3

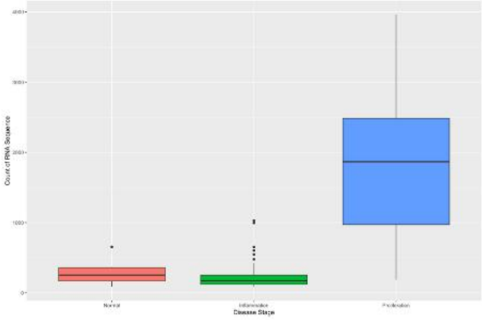

TMEM256

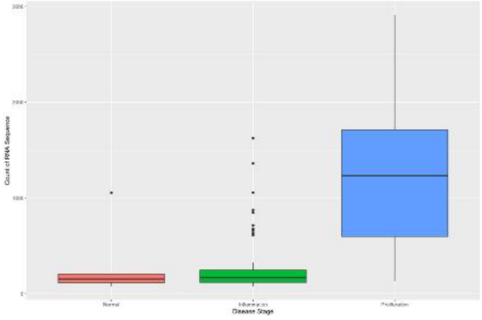

TUBA1B

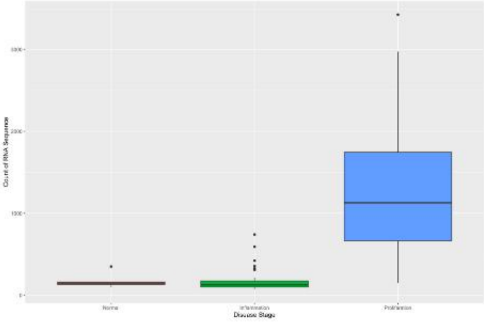

VBP1

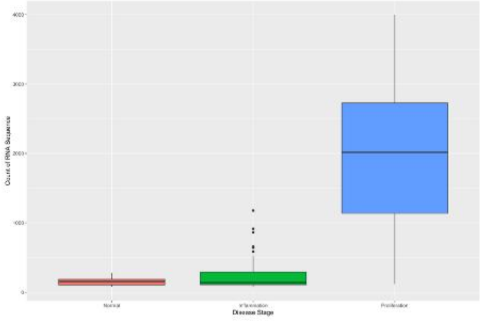

YTHDC1

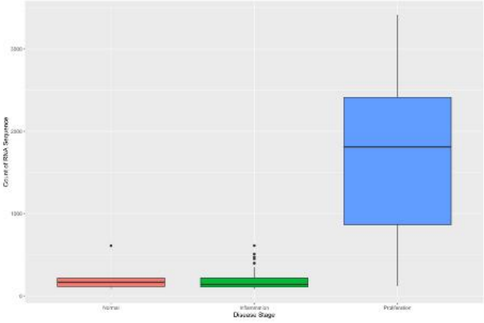

ZFP36L1
